# Effectiveness of interventions targeting air travellers for delaying local outbreaks of SARS-CoV-2

**DOI:** 10.1101/2020.02.12.20022426

**Authors:** Samuel Clifford, Carl A.B. Pearson, Petra Klepac, Kevin Van Zandvoort, Billy J. Quilty, CMMID COVID-19 working group, Rosalind M. Eggo, Stefan Flasche

**Affiliations:** Centre for Mathematical Modelling of Infectious Diseases, Department of Infectious Disease Epidemiology, London School of Hygiene and Tropical Medicine, Keppel Street, WC1E 7HT London, UK

## Abstract

**Background:** We evaluated if interventions aimed at air travellers can delay local SARS-CoV-2 community transmission in a previously unaffected country.

**Methods:** We simulated infected air travellers arriving into countries with no sustained SARS-CoV-2 transmission or other introduction routes from affected regions. We assessed the effectiveness of syndromic screening at departure and/or arrival & traveller sensitisation to the COVID-2019-like symptoms with the aim to trigger rapid self-isolation and reporting on symptom onset to enable contact tracing. We assumed that syndromic screening would reduce the number of infected arrivals and that traveller sensitisation reduces the average number of secondary cases. We use stochastic simulations to account for uncertainty in both arrival and secondary infections rates, and present sensitivity analyses on arrival rates of infected travellers and the effectiveness of traveller sensitisation. We report the median expected delay achievable in each scenario and an inner 50% interval.

**Results:** Under baseline assumptions, introducing exit and entry screening in combination with traveller sensitisation can delay a local SARS-CoV-2 outbreak by 8 days (50% interval: 3-14 days) when the rate of importation is 1 infected traveller per week at time of introduction. The additional benefit of entry screening is small if exit screening is effective: the combination of only exit screening and traveller sensitisation can delay an outbreak by 7 days (50% interval: 2-13 days). In the absence of screening, with less effective sensitisation, or a higher rate of importation, these delays shrink rapidly to less than 4 days.

**Conclusion:** Syndromic screening and traveller sensitisation in combination may have marginally delayed SARS-CoV-2 outbreaks in unaffected countries.

## Background

Similar to outbreaks of other respiratory pathogens (1–4), syndromic airport screening at arrival of travellers from regions with a high risk of SARS-CoV-2 infection is unlikely to identify a sufficient proportion of infected travellers to prevent global spread (5,6). Sensitising arriving travellers to the symptoms and risk of SARS-CoV-2 and encouraging appropriate reactions (e.g., early self-isolation, requesting medical assistance via telephone, reporting travel history to providers to trigger tracing and quarantine of contacts), may have a more pronounced effect and has been implemented in many transport hubs (7). Unfortunately, with increasing numbers of infected travellers contact tracing is unlikely to be sustainable for long because of the immensely resource-intensive nature of contact tracing and hence is similarly unlikely to prevent local transmission in the long term (8).

Even if containment is ultimately impossible, delaying local spread remains a key target of pandemic response (9). This will allow additional time for preparation of the health system and mobilisation of additional public health resources. Delaying local spread will also allow for crucial time to better understand the pathogen and to evaluate effective treatment and prevention measures.

We aim to estimate the effectiveness of syndromic screening and traveller sensitisation for delaying the onset of sustained SARS-CoV-2 spread in previously unaffected regions.

## Methods

### Infected Traveller Arrivals Model

We represent the potential importation of infections by a non-homogeneous Poisson process with intensity function, *λ*(*t*), representing the instantaneous rate of arrival of infected travellers (per week), and that the travellers are attempting to travel to a specific country or region currently not experiencing an outbreak. Implicitly, the number of infected travellers is a product of the prevalence and the number of travellers per week. We assume that individuals with severe symptoms do not attempt to travel, though travellers may develop severe symptoms en route (6).

For early stages of an outbreak, with sufficient control measures in place at the source of the outbreak to flatten or reverse the spread, it may be reasonable to assume a constant arrival rate. Instead we assume that *λ*(*t*) grows from an initial rate, *λ*_0_infected travellers per week, when measures to limit the spread from imported cases are introduced. The assumed exponential growth rate of *r* = 0.1 (95% CI, 0.050-0.16) corresponds to an epidemic doubling time of 7.4 days (95% interval: 4.2-14 days), in line with the local epidemic growth during attempted control via contact tracing but without a lockdown (10). We consider that the epidemic grows exponentially at the source during the early phase of the outbreak when the population is effectively entirely susceptible. In addition to their use in modelling the risk of exportation of SARS-CoV-2 (11) and turning points for daily case trends in SARS-CoV (12), non-homogeneous Poisson processes, particularly those with decreasing inter-arrival times, have previously been applied to a range of infectious disease settings for investigating the effectiveness of border control (13), estimating epidemic parameters (14) and assessing scheduling in mass immunisation clinics for pandemic Influenza (15).

### Outbreak Probability Model

Upon arrival, we assume all infected travellers have the same distribution of the number of onwards infections they would generate if circulating in the community. These potential secondary infections are determined by the average number of those infections, *R*_0_, the basic reproduction number, and the dispersion of that number, *k*.

Following Hartfield and Alizon (16), we calculate the probability that an arriving infected traveller causes an outbreak, given *R*_0_ and dispersion parameter,*k*, by solving the first equation in their Supplementary Material S.3,

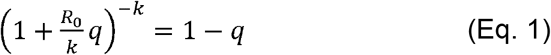

for *q*, the probability that an outbreak is triggered by an arriving traveller (Hartfield and Alizon parameterise this in terms of *s* = 1 − *q* the probability of extinction of an outbreak).

Having obtained *q* for a given simulation we calculate *N*_0_, the number of infected travellers required to trigger the outbreak from a geometric distribution with probability *q*, sampling the *u* ∼ *U*(0,1) quantile of said distribution to match initial conditions between intervention scenarios across simulation samples. We assume that the arrival times of infected travellers follows a non-homogeneous Poisson process with intensity *λ*(*t*) = *λ* _0_ *e*^*rt*^ /7, where *λ* _0_/7 is the arrival rate of infected travellers (per day) when the interventions are introduced at *t* = 0 and the rate of increase, *r*, is sampled from a Gamma distribution with 95% interval (0.05, 0.16) representing the growth early in the Wuhan outbreak (10). Additionally, because *R*_0_ is uncertain, this leads to uncertainty in *q* and variability in *t*_0_ beyond the stochasticity from the arrival process. This exponential growth rate is consistent with a case doubling times of: 5 days (95% interval: 4.3, 6.2 days) found by Ferretti et al. (17) which would give a growth rate of approximately 0.14; and that of 6.4 days (5.8, 7.1 days) by Wu et al. (18) giving a growth rate of 0.11 days. We have chosen to parameterise in terms of the exponential growth rate of the epidemic rather than the doubling time of cases to account for recovery at the travel origin.

### Model of symptom screening and sensitisation

When syndromic screening is implemented, each arriving infected traveller is identified during screening with probability 1 − *θ*, reducing the number of infected travellers arriving and potentially delaying the outbreak. For the scenarios we consider, we assume the same baseline assumptions as in Quilty et al (6); i.e. a syndromic screening sensitivity of 86%, travel duration of 12 hours, and average times from infection to onset of symptoms and from onset to severe symptoms/hospitalisation as 5.2 and 9.2 days, respectively. For those assumptions, Quilty et al estimate the mean probability of SARS-CoV-2 infected travellers not being detected at either exit or entry screening as 46% and as 42% for exit-only screening. Here we consider the uncertainty in *θ* by bootstrap resampling 100 travellers per simulation from the model of Quilty et al. and obtain 95% confidence intervals of (33%, 53%) and (37%, 57%) respectively. As in that paper, the benefit of entry screening is dependent on the effectiveness of exit screening, and entry-only screening is likely to pick up those who would have been identified by exit screening.

Sensitisation occurs via, e.g., posters and handouts to travellers arriving from high risk regions, which increases the likelihood that those travellers, if they experience SARS-CoV-2 symptoms, will self-isolate on the occurrence of mild symptoms and rapidly report to health care providers who in turn trigger contact tracing (8). We represent traveller sensitisation as reducing *R*_0_ to *R*′ = (1 − *ϱ*)*R*_0_, where *ϱ* is interpreted as the effectiveness of sensitisation, rather than the proportion of passengers perfectly sensitised. The lower *R*′ results in a lower probability that an arriving infected traveller triggers an outbreak, *q*′, and therefore it may require the entry of more infected travellers, *N*′ ≥ *N*, to trigger the outbreak than in the no-sensitisation case, resulting in a correspondingly longer time to outbreak in the Poisson process.

As a base case for the intervention, we consider recent work (5) which indicates that sensitisation by itself may cause only 25% of those symptomatically infected with SARS-CoV-2 to self-report upon onset of symptoms. In line with Hellewell et al (8) we assume, for sensitivity, a best case scenario that these measures accelerate self-isolation and reporting in the early stages of the SARS-CoV-2 pandemic and reduce the average number of onward transmitting secondary infections by about 50%.

### Calculation of delays to reach outbreak threshold

To determine the impact of the interventions, we calculate the difference in time to outbreak occurrence with and without interventions: *t*′ − *t*_0_. For the correct comparison, these times must be drawn as matched quantiles, *u* ∼ *U*(0,1). To ensure that, we calculate *N*_*0*_ and *N*′ from the same *R*_0_ draw (reduced to *R*′ by sensitisation and contact tracing) and calculate the probability of an individual traveller causing an outbreak without and with interventions, *q* and *q*′, respectively. We then generate arrival times from Poisson processes with rate *λ*(*t*) and determine how long it takes for *N*_*0*_ infected travellers to arrive in the base case and *N*′ to arrive in the screened queue. Arrival times are generated using the reda package (19) in R 3.6.2.

The expected arrival day for the *N*th infected traveller in the no-intervention case, given *r, λ*_0_ is calculated by integrating the exponentially increasing intensity, 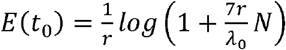. The expected arrival day of the *N*′ th infected traveller under the intervention is 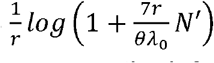 Where *R*′ is less than 1, *N*′ is infinite and the simulated outbreak does not occur as the infected traveller causing the outbreak will never arrive.

We report the median delay and inner 50% and 95% intervals and plot the empirical complementary cumulative probability densities to show how many simulations result in a delay of at least a given duration given *k, λ*_*0*_, *ϱ, θ*.

### Scenarios considered

We considered three syndromic screening intervention scenarios: no screening, exit-only, and exit-and-entry screening. We further considered two scenarios of the effectiveness of traveller sensitisation: 0% and a reasonable average case of 25%. No screening and 0% sensitisation effectiveness form the non-intervention reference. These are reported in the context of either 0.1, 1, 10 or 100 infected travellers per week at the time of measures being introduced. We assume that the mean *R*_0_is gamma distributed with an inner 95% range from 1.4 to 3.9 (10); we assume, for the calculation of probability of outbreak triggering, the dispersion in secondary cases is k=0.54 (20).

For sensitivity analyses, we also investigate alternative scenarios for the dispersion of *R*_0_and a reasonable best case of 50% effectiveness of contact tracing and self-reporting. All scenarios and parameters are summarised in Table 1.

**Table 1:**
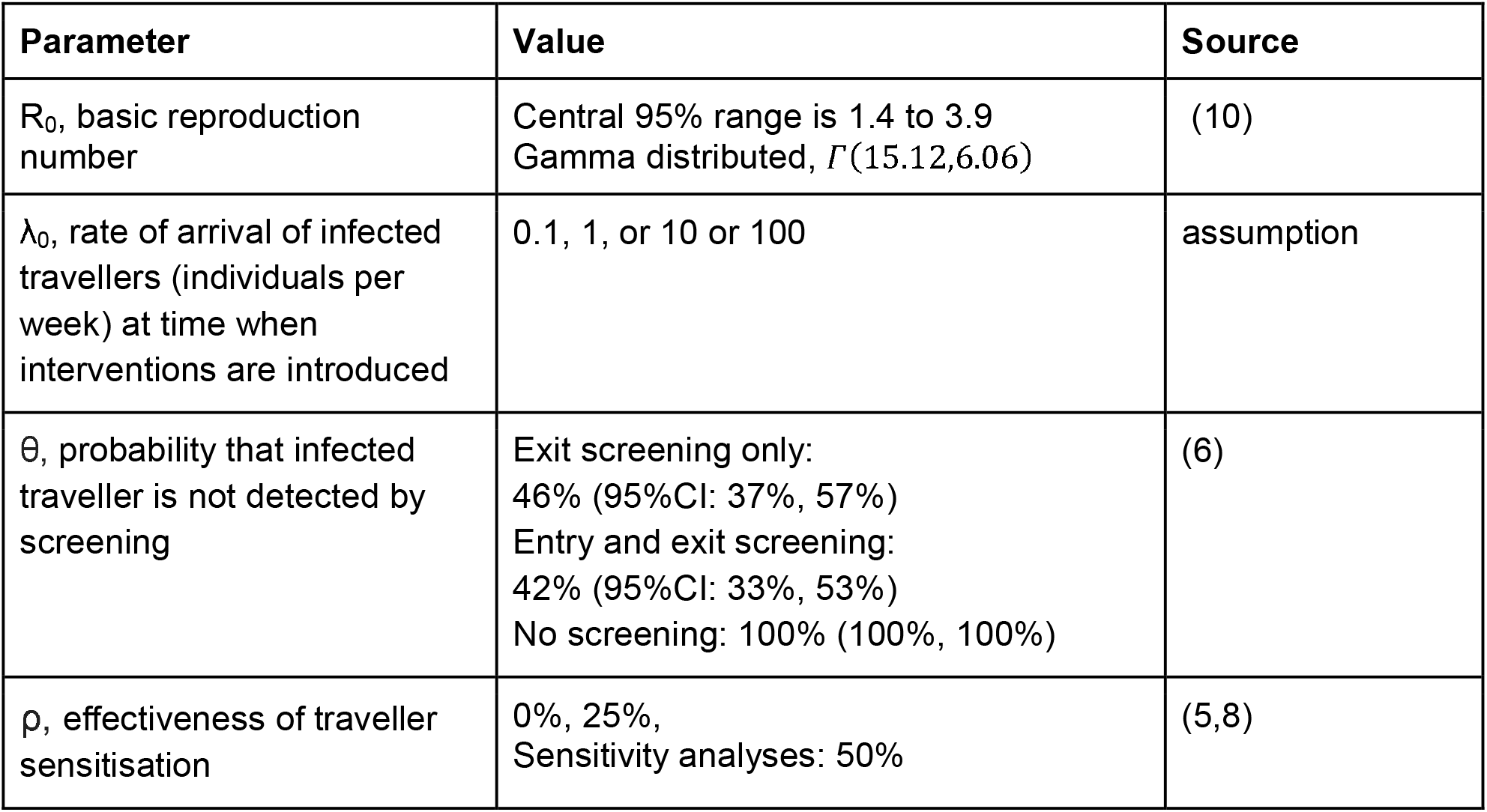
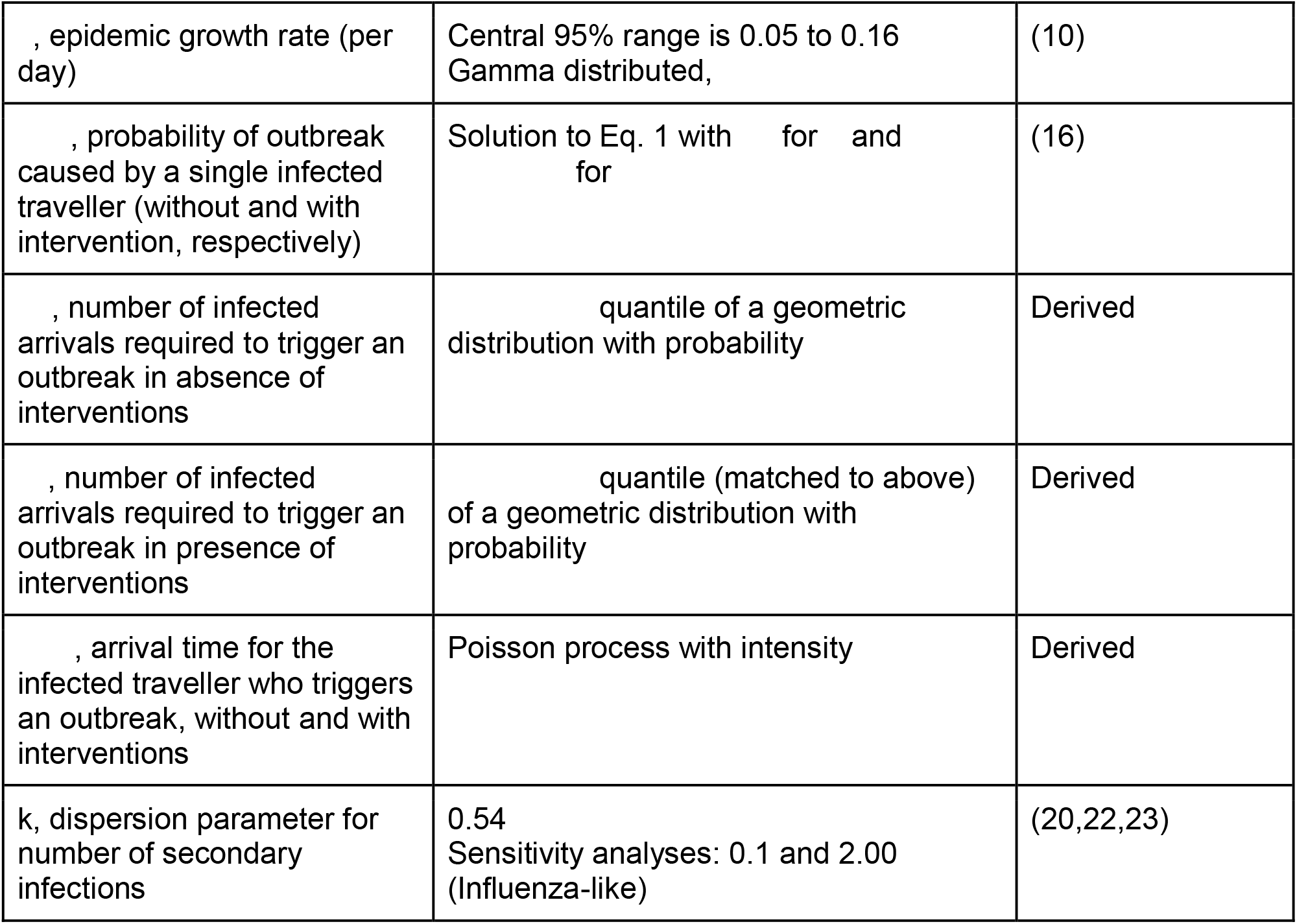
Overview of parameter assumptions for the model.

| Parameter | Value | Source |
| --- | --- | --- |
| $R_0$, basic reproduction number | Central 95% range is 1.4 to 3.9<br>Gamma distributed, $\Gamma(15.12, 6.06)$ | (10) |
| $\lambda_0$, rate of arrival of infected travellers (individuals per week) at time when interventions are introduced | 0.1, 1, or 10 or 100 | assumption |
| $\theta$, probability that infected traveller is not detected by screening | Exit screening only:<br>46% (95%CI: 37%, 57%)<br>Entry and exit screening:<br>42% (95%CI: 33%, 53%)<br>No screening: 100% (100%, 100%) | (6) |
| $\rho$, effectiveness of traveller sensitisation | 0%, 25%,<br>Sensitivity analyses: 50% | (5,8) |
| , epidemic growth rate (per day) | Central 95% range is 0.05 to 0.16<br>Gamma distributed, | (10) |
| , probability of outbreak caused by a single infected traveller (without and with intervention, respectively) | Solution to Eq. 1 with for and for | (16) |
| , number of infected arrivals required to trigger an outbreak in absence of interventions | quantile of a geometric distribution with probability | Derived |
| , number of infected arrivals required to trigger an outbreak in presence of interventions | quantile (matched to above) of a geometric distribution with probability | Derived |
| , arrival time for the infected traveller who triggers an outbreak, without and with interventions | Poisson process with intensity | Derived |
| k, dispersion parameter for number of secondary infections | 0.54<br>Sensitivity analyses: 0.1 and 2.00 (Influenza-like) | (20,22,23) |

All analyses were done with R 3.6.2 (21) and can be found on GitHub at https://github.com/cmmid/screening_outbreak_delay/.

**Figure 1:**
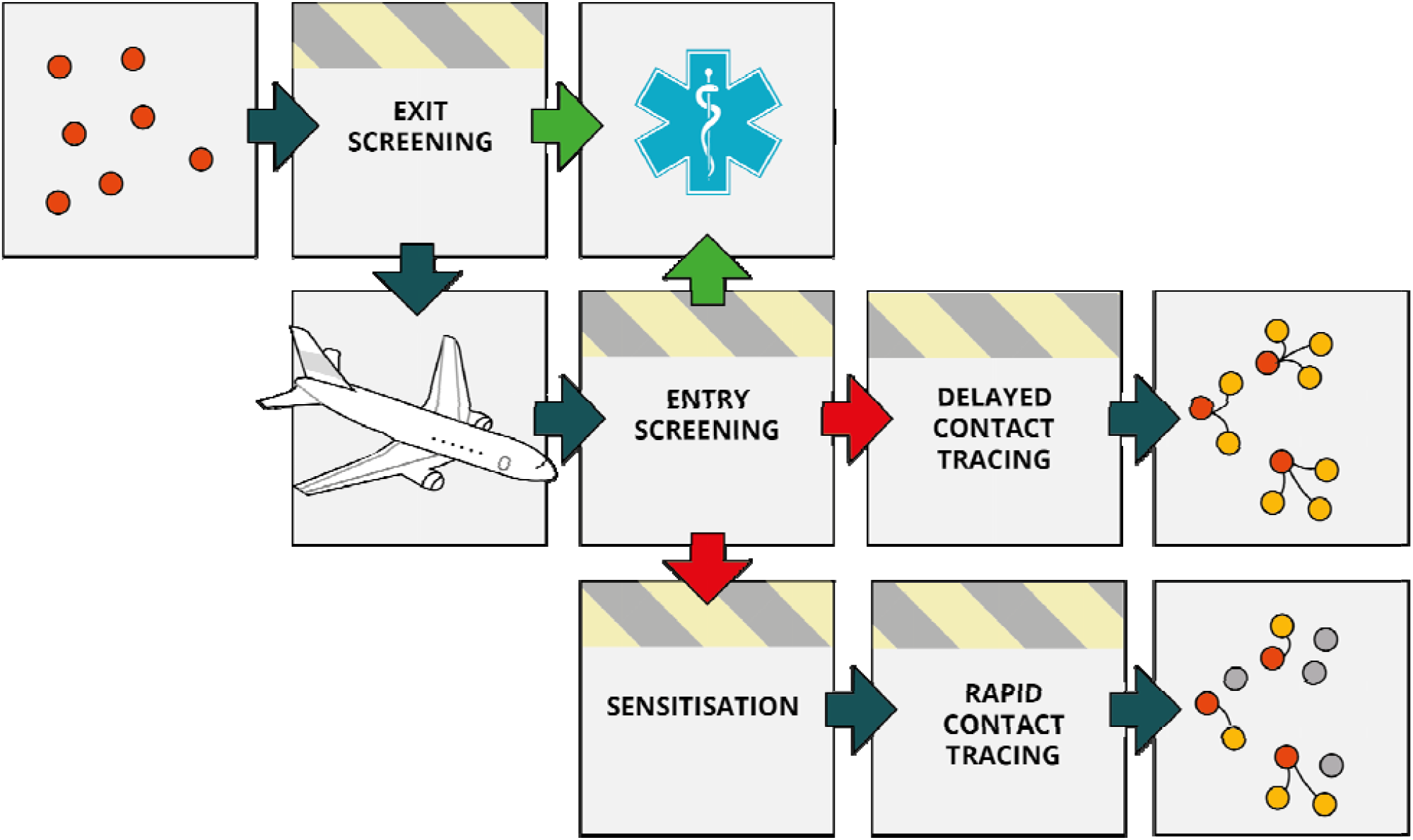
Schematic of the air traveller intervention process. A proportion of infected travellers (red dots) will be detected through syndromic exit or entry screening (green arrows) and will immediately be isolated and not cause secondary cases (yellow dots) in the yet unaffected destination. Travellers not identified by syndromic screening enter the destination country (red arrows), where they are provided by sensitisation information and are more likely to self-isolate and/or report their symptoms soon after onset and cause fewer secondary cases (dots which are yellow under “delayed contact tracing” but grey under “rapid contact tracing”).

## Results

For all scenarios investigated, the lower bound of the 95% interval is always less than 1 day of delay (Table 2). Where sensitisation has been performed (*ϱ* either 25% or 50%), the sampled value of *R*′ may be less than 1; for such values, the outbreak is averted. Where the upper bound of the 95% interval is infinite, this indicates that at least 2.5% of outbreaks have been averted. Here we present results in terms of their median and inner 50% interval and only present the upper bounds of the 95% interval when they are finite. The percentage of outbreaks averted for all combinations of *ϱ* and *θ* are given in Table S1.

**Table 2:** Summary statistics providing the inner 50% and 95% confidence intervals and medians (all rounded to the nearest day) for the estimated number of days an outbreak is delayed given one arriving infection per week at the introduction of an intervention consisting of a combination of traveller screening and sensitisation and contact tracing. Comparisons are made to no contact tracing and no screening (there are no “No screening” results at 0% sensitisation as this is the baseline against which comparisons are to be made).

| Arrivals/week, $\lambda$ | Sensitisation, $\varrho$ | Screening | Number of days for which the given percentage of delays are at least this long | | | | |
| --- | --- | --- | --- | --- | --- | --- | --- |
|  |  |  | 97.5% | 75% | 50% | 25% | 2.5% |
| 0.1 | 0% | Exit and entry | <1 | <1 | 6 | 13 | 41 |
|  |  | Exit only | <1 | <1 | 5 | 12 | 41 |
| | 25% | Exit and entry | <1 | 4 | 12 | 22 | ∞ |
| | | Exit only | <1 | 3 | 10 | 20 | ∞ |
| | | No screening | <1 | <1 | 1 | 7 | ∞ |
| 1 | 0% | Exit and entry | <1 | <1 | 4 | 9 | 23 |
|  |  | Exit only | <1 | <1 | 3 | 8 | 21 |
| | 25% | Exit and entry | <1 | 3 | 8 | 14 | ∞ |
| | | Exit only | <1 | 2 | 7 | 13 | ∞ |
| | | No screening | <1 | <1 | 1 | 4 | ∞ |
| 10 | 0% | Exit and entry | <1 | <1 | 1 | 3 | 8 |
|  |  | Exit only | <1 | <1 | 1 | 2 | 7 |
| | 25% | Exit and entry | <1 | <1 | 2 | 5 | ∞ |
| | | Exit only | <1 | <1 | 2 | 4 | ∞ |
| | | No screening | <1 | <1 | <1 | 1 | ∞ |
| 100 | 0% | Exit and entry | <1 | <1 | <1 | <1 | 1 |
|  |  | Exit only | <1 | <1 | <1 | <1 | 1 |
| | 25% | Exit and entry | <1 | <1 | <1 | 1 | ∞ |
| | | Exit only | <1 | <1 | <1 | 1 | ∞ |
| | | No screening | <1 | <1 | <1 | <1 | ∞ |

In the case of 1 infected traveller per week at the time of the intervention, the combination of traveller sensitisation and exit and entry screening typically delays the outbreak by 8 days (inner 50% interval: 3-14 days) (Table 2, Figure 2). If there are 10 infected travellers per week at the time of these interventions being introduced, the outbreak is typically delayed by only 2 days (50%: <1-5 days). At *λ*_0_ = 100the median delay is less than 1 day, and less than 25% of delays are longer than 1 day.

Additional figures in the appendix show the complementary cumulative density functions, focusing on either variation with screening (Figure S1), traveller sensitisation (Figure S2), arrival rate (Figure S3), or dispersion parameter (Figure S4).

The incremental benefit of syndromic entry screening is highly dependent on the effectiveness of exit screening. With one infected traveller per week, traveller sensitisation, and under baseline assumptions of exit screening effectiveness but no entry screening, the outbreak is delayed by 2 days (50%: <1-13 days), indicating that additional entry screening adds little in this case.

We estimate that with no traveller sensitisation and under baseline assumptions for the effectiveness of syndromic screening at exit and entry, the delays are half as long as if the effect of sensitisation was 25%. In the early stages of the outbreak with 1 infected traveller per week at the time the intervention is introduced, an outbreak may be delayed by screening alone by only 4 days (50%: <1-9 days). Again, this is largely due to exit screening at departure, which on its own is estimated to delay the outbreak by 3 days (50%: <1-8 days). Forgoing screening measures until a rate of arrival of 10 infected travellers per week, essentially eliminates any potential delay in onset of a local outbreak; specifically, if infected traveller numbers approach 10 per week, syndromic screening alone can only delay the outbreak by 1 day (50%: <1-3 days).

Similarly, we estimate that in the absence of syndromic air traveller screening, traveller sensitisation can only delay the outbreak by 1 day (50%:<1-4 days) early in the epidemic when the arrival rate is 1 infected traveller per week at time of introduction of the intervention. When the rate of arrival at time of introducing only sensitisation is 10 infected travellers per week, the introduction of sensitisation results in delays of less than 1 day (50%: <1-1 day) and that at 100 infected arrivals per week there is no delay unless the outbreak is completely averted (only the case for 13% of simulated outbreaks).

For sensitivity analyses, we varied the effectiveness of traveller sensitisation and the heterogeneity in the number of secondary infections. A 50% reduction in the effective reproduction number through traveller sensitisation followed by rapid case isolation and contact tracing can potentially prevent a local outbreak independent of the number of infected arrivals if the basic reproduction number is smaller than 3.3 (i.e., *R*′ = *R*_0_ (1 − *ϱ*) < 1). As traveller sensitisation increases and therefore a greater proportion of simulated *R*′ values are less than 1, the proportion of simulated delays that are infinitely long (indicating that that specific simulated potential outbreak has been averted) increases to nearly 66% (Table S1).

If the number of secondary infections is substantially less disperse, e.g. influenza-like, fewer outbreaks are averted, and median outbreak delays decrease by about 25%, as the outbreak becomes less reliant on occasional super-spreading events (Figure S4). If, however, the number of secondary infections is slightly more disperse, i.e. the dispersion parameter estimate of 0.1 (23), then outbreak delays are also approximately 25% shorter but fewer outbreaks are averteed than in the *k =* 2 case (Table S1).

**Figure 2:**
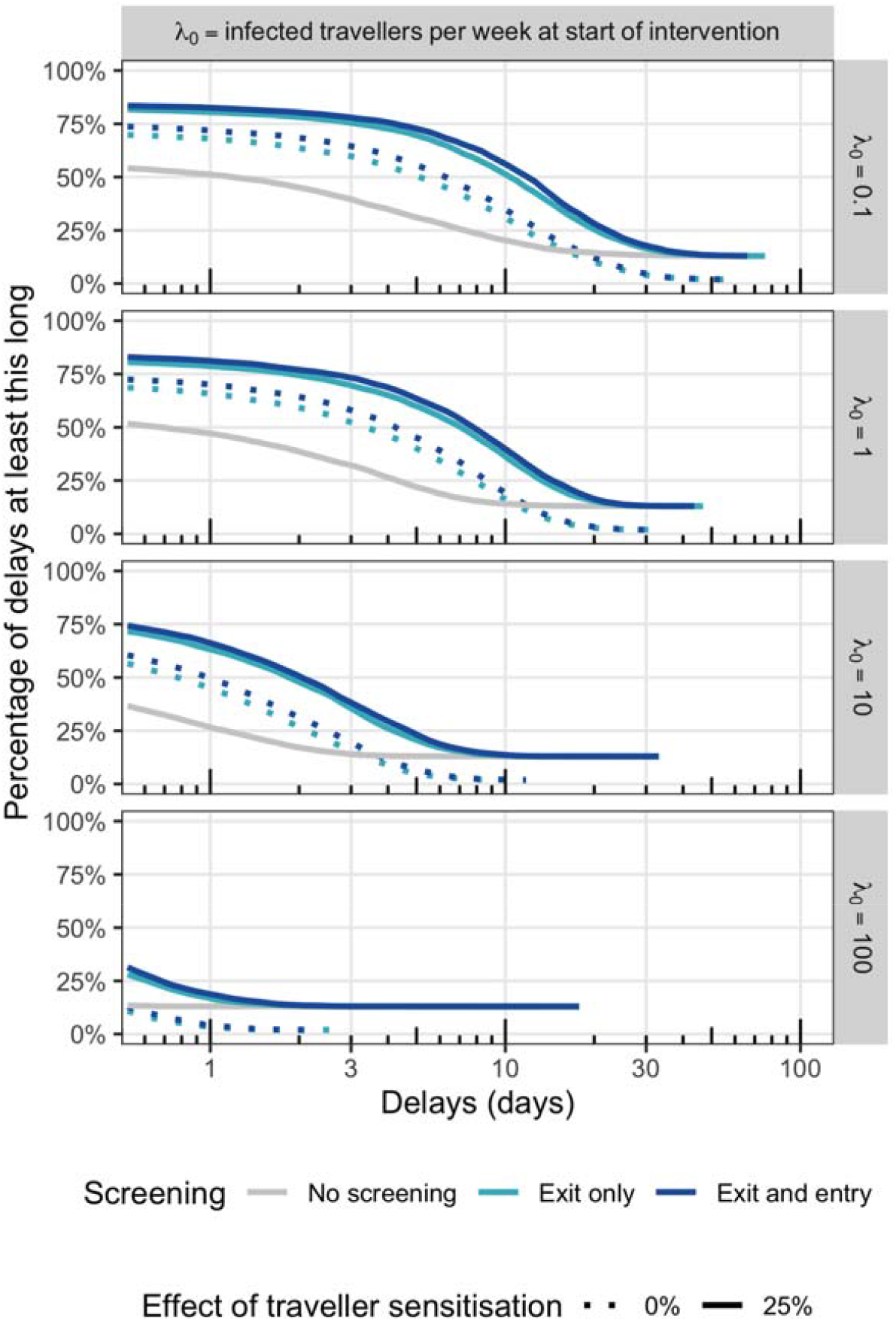
Complementary empirical cumulative density functions for the estimated number of days an outbreak is delayed given an intervention consisting of a combination of traveller screening and sensitisation and contact tracing. Rows correspond to different arrival rates and columns to traveller sensitisation. Comparisons are made to no contact tracing and no screening (there are no “No screening” results at 0% sensitisation as this is the baseline against which comparisons are to be made).

## Discussion

Syndromic screening of air travellers at departure and/or arrival is unlikely to prevent a sufficient proportion of SARS-CoV-2 infected travellers from entering a yet unaffected country and thereby prevent a local outbreak. Similarly, sensitisation of travellers from high-risk countries to encourage self-isolation and enable accelerated case detection and contact tracing if indeed infected will likely not be able to halt an outbreak indefinitely, particularly when many infected travellers arrive undetected, unless the effect of sensitisation is large enough ensure that the number of secondary infections are, on average, less than 1 for the traveller and subsequent cases. We investigate here how syndromic screening and traveller sensitisation, as well as their combination, may delay an outbreak of SARS-CoV-2.

We find that when syndromic screening alone is introduced very early in the outbreak, i.e. at a point when 0.1 infected travellers per week arrive, it can only slightly delay an outbreak (6 days, 50%: <1-13 days). Traveller sensitisation alone has a less pronounced effect by delaying the outbreak by 1 day (50%: <1-7 days). The combination of syndromic screening and traveller sensitisation may more substantially delay an outbreak while the number of infected travellers is this low (12 days, 50%: 4-22). The incremental effect of syndromic entry screening is only notable if exit screening is poor or even absent. These results are sensitive to a number of key assumptions: with increasing R_0_, less heterogeneous R_0_ or less effective traveller sensitisation the estimated achievable delay quickly becomes negligible. Furthermore, once the weekly number of infected passengers increases to 10 and above, e.g. as a result of an exponential increase in cases at the origin of travel, even the combination of syndromic screening and traveller sensitisation delays is unlikely to delay an outbreak for more than a week.

We find a potential small role for interventions targeting air-passengers to delay major outbreaks of SARS-CoV-2 in previously unaffected regions as long as implemented very early in an outbreak. We find that syndromic screening on arrival can add to the effect of traveller sensitisation in these early stages of a pandemic. Syndromic screening can also aid to reduce the number of passengers that would eventually self-report and then require resource-intensive follow up, including contact tracing. As the rate of infected arrivals increases, contact tracing becomes increasingly more difficult and the effectiveness is likely to decrease, further shortening the achievable delays. Therefore, syndromic screening may have an additional role in helping to sustain control efforts for longer. Of note, however, is that syndromic screening at arrival only substantially adds to control efforts if syndromic screening at departure is absent or largely ineffective.

Delays in airport processes arising from screening may expose travellers to additional risk depending on airport design and reduction in pedestrian flow rates within the terminal and therefore the amount of time passengers spend waiting in crowded areas (24) as well as the time spent boarding and alighting (25). While not as long in duration as the flight itself, during their time in terminals, travellers mix with a much larger and more diverse range of people than during the flight. This is outside the scope of this study, however, and relies on assumptions about background prevalence in the community of airport users and mixing within airports.

While our findings may encourage implementation of both syndromic screening on entry and traveller sensitisation in the early stages of the SARS-CoV-2 pandemic, it is important to note that these findings are highly sensitive to the underlying base-case assumptions and do not consider the economic implications of large scale air passenger screening and contact tracing (26). Despite the cost, however, the argument could be made that public health (which enables ongoing economic health) is more important a goal during a pandemic than short-term budgetary considerations (27) particularly in the absence of a vaccine.

Wells et al. (28) focus on the risk of exporting, rather than importing, the virus and estimate the risk of exportation from China to another country given weights based on airline movements and distributions of incubation time. They considered the impact of travel and border restrictions and found that these restrictions decreased the daily rate of exportation from mainland China to other countries by 81% in the three weeks after introduction, and averted 71% of the cases that they estimated would have occurred had no lockdown been introduced. This would provide countries without established outbreaks to take measures to further delay, e.g. screening, sensitisation and contact tracing, as well as preparing their health systems for the outbreak (7,29).

With increasing numbers of infected travellers, a higher number of secondary infections or a lower heterogeneity thereof, or less effective interventions, the achievable delay quickly drops down to a few days of delay. While all of our assumptions include the best knowledge on SARS-CoV-2 to date, there is considerable uncertainty associated with all of these assumptions. For example, we have assumed recently reported heterogeneity in the individual R_0_, however, the reported range of uncertainty includes SARS-like and influenza-like which can drastically alter the results. Some recent estimates would suggest more SARS-like or even more overdispersed *k* which would imply that longer outbreak delays are possible as shown in our sensitivity analysis (30). We also don’t explicitly account for potential asymptomatic transmission. However, we implicitly do so as both the syndromic screening as well as the contact tracing work that informed our estimates accounted for a small proportion of asymptomatic transmitters who we assume similarly transmit but will not be affected by syndromic screening or sensitisation. Furthermore, the results are predicated on a syndromic screening sensitivity of 86% (6). When reducing the sensitivity to 70%, as used in other reports (5), delays reduce by about 20%.

Travel restrictions were implemented in the form of flights exiting China being suspended (31), which has potentially curbed the exponential increase of infected travellers despite an exponential increase of infections in China. Assuming exponential growth with *r* = 0.1numbers of infected arrivals would have increased from 1 to 10 and 100 per week within about 23 and 46 days respectively (assuming exponential growth with *r* = 0.1) and estimated delays would decrease accordingly. However, infected traveller arrivals likely still have increased exponentially as a result of the largely undetected spread in Iran and Italy (32) early on in the pandemic (increasing, respectively, from 28 and 76 cases as of 23 Feb. 2020 (33) to 593 and 1128 a week later on 1 Mar. 2020 (34)).

By February, many major airlines had suspended flights from mainland China with travel restrictions from Iran, Italy and Korea being added more recently. In the three weeks leading up to the 28th February the UK reported 10 imported cases, 4 of them in the final week (35). At that early point a more optimistic scenario would have been that the control measures in place limit the number of infected travellers and may sustainably do so for a considerable amount of time. This constant rate of importation, which is more similar to e.g. SARS in 2003 would have led to much larger possible delays in local outbreak through targeting of air travellers.

Future pandemic threats will bear similar questions. While our considerations are focussed around SARS-CoV-2 prevention there are some generally applicable conclusions. The expected delay of a local outbreak as a result of traveller targeted interventions will depend on the pathogen specific epidemiology but potential pre- and asymptomatic transmission are a key challenge to the success of such. Further, for pathogens with long incubation period, syndromic screening is likely to miss many infected passengers and a high reproduction number increases the chance that a single missed infected will trigger a local outbreak.

In summary, we find that the targeting of air-travellers with syndromic screening at exit or entry and sensitisation for signs of symptoms following their arrival has likely delayed the local spread of SARS-CoV-2, but only by a few days. This is because measures were largely put in place at a time where already a few infected travellers a week were arriving and that infection prevalence among travellers was likely increasing exponentially. We find that syndromic screening at arrival may enhance control efforts, but only in the absence of syndromic screening at departure.

## Data Availability

The study contains no primary data

https://github.com/samclifford/screening_outbreak_delay/

## Acknowledgements

SF and SC are supported by a Sir Henry Dale Fellowship jointly funded by the Wellcome Trust and the Royal Society (Grant number 208812/Z/17/Z). RME acknowledges an HDR UK Innovation Fellowship (Grant number MR/S003975/1). BQ was funded by the National Institute for Health Research (NIHR) (16/137/109) using UK aid from the UK Government to support global health research. PK was funded by the Bill & Melinda Gates Foundation (INV-003174). KvZ, is supported by Elrha’s Research for Health in Humanitarian Crises (R2HC) Programme, which aims to improve health outcomes by strengthening the evidence base for public health interventions in humanitarian crises. The R2HC programme is funded by the UK Government (DFID), the Wellcome Trust, and the UK National Institute for Health Research (NIHR). The views expressed in this publication are those of the author(s) and not necessarily those of the NHS, the NIHR or the UK Department of Health and Social Care. The following authors were part of the Centre for Mathematical Modelling of Infectious Disease SARS-CoV-2 working group and contributed equally and appear in random order: Christopher I Jarvis, Yang Liu, Nikos I Bosse, Kiesha Prem, Adam J Kucharski, W John Edmunds, Timothy W Russell, Sebastian Funk, Mark Jit, Hamish Gibbs, Sam Abbott, James D Munday, Amy Gimma, Nicholas Davies, Charlie Diamond, Joel Hellewell. Each contributed in processing, cleaning and interpretation of data, interpreted findings, contributed to the manuscript, and approved the work for publication. We also like to thank John Edmunds, Graham Medley and Annelies Wilder-Smith for their helpful comments during the conception of this work.

Neither patients nor the public were involved with the design, conduct, reporting, or dissemination plans of our research. As this work is a simulation study, there are no participants to which we can disseminate the results of this research.

## Appendix

**Figure S1:**
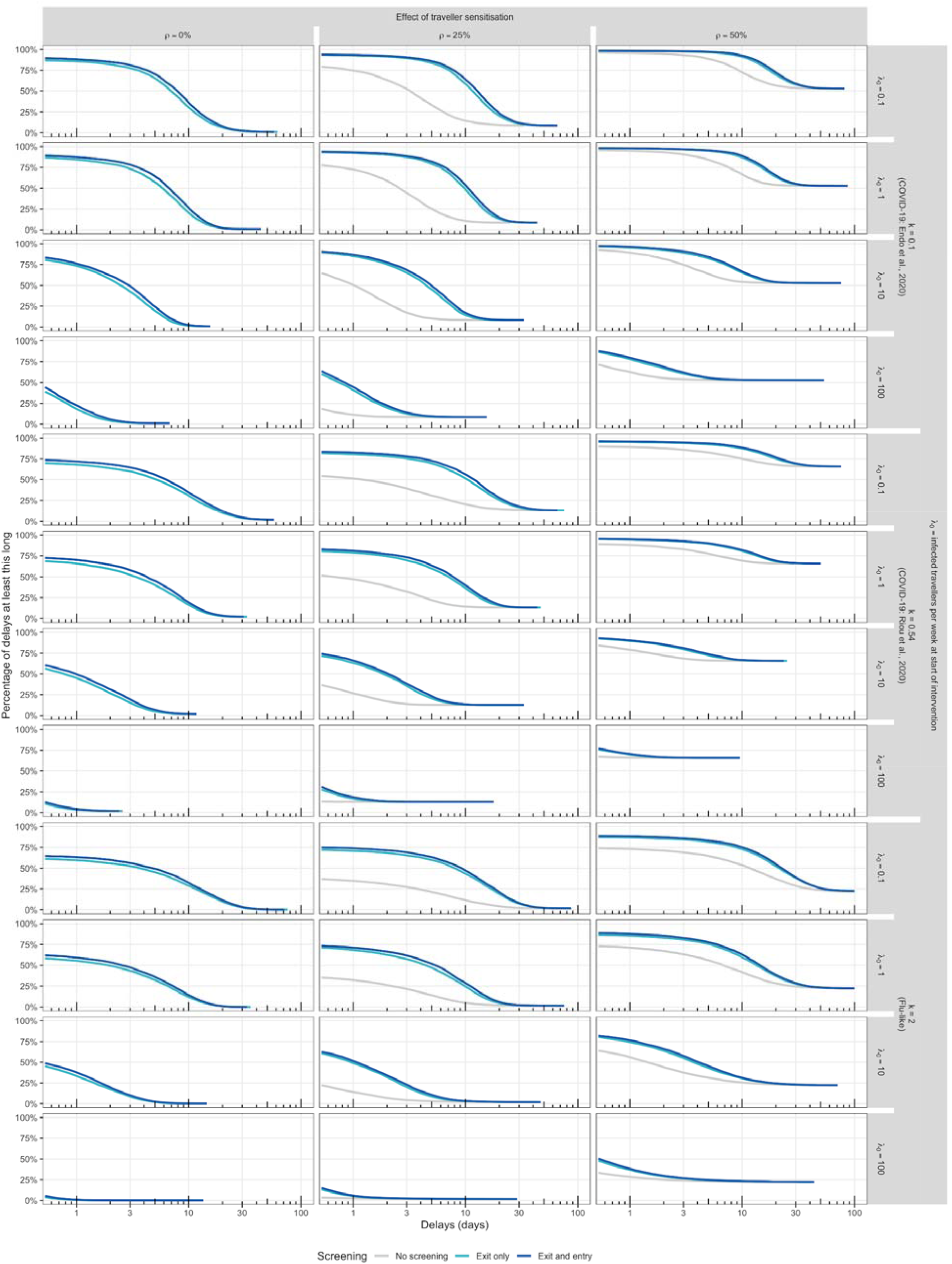
All scenarios for Figure 2 - complementary cumulative density functions (CCDF = 1-ECDF) of the estimated number of days an outbreak is delayed given an intervention consisting of a combination of traveller screening and sensitization and contact tracing. Within each panel, and for a given delay, the CCDF shows the percentage of simulations which result in a delay of at least that long for each screening regime (no screening, exit only, exit and entry). Comparisons are made to no contact tracing and no screening.

**Figure S2:**
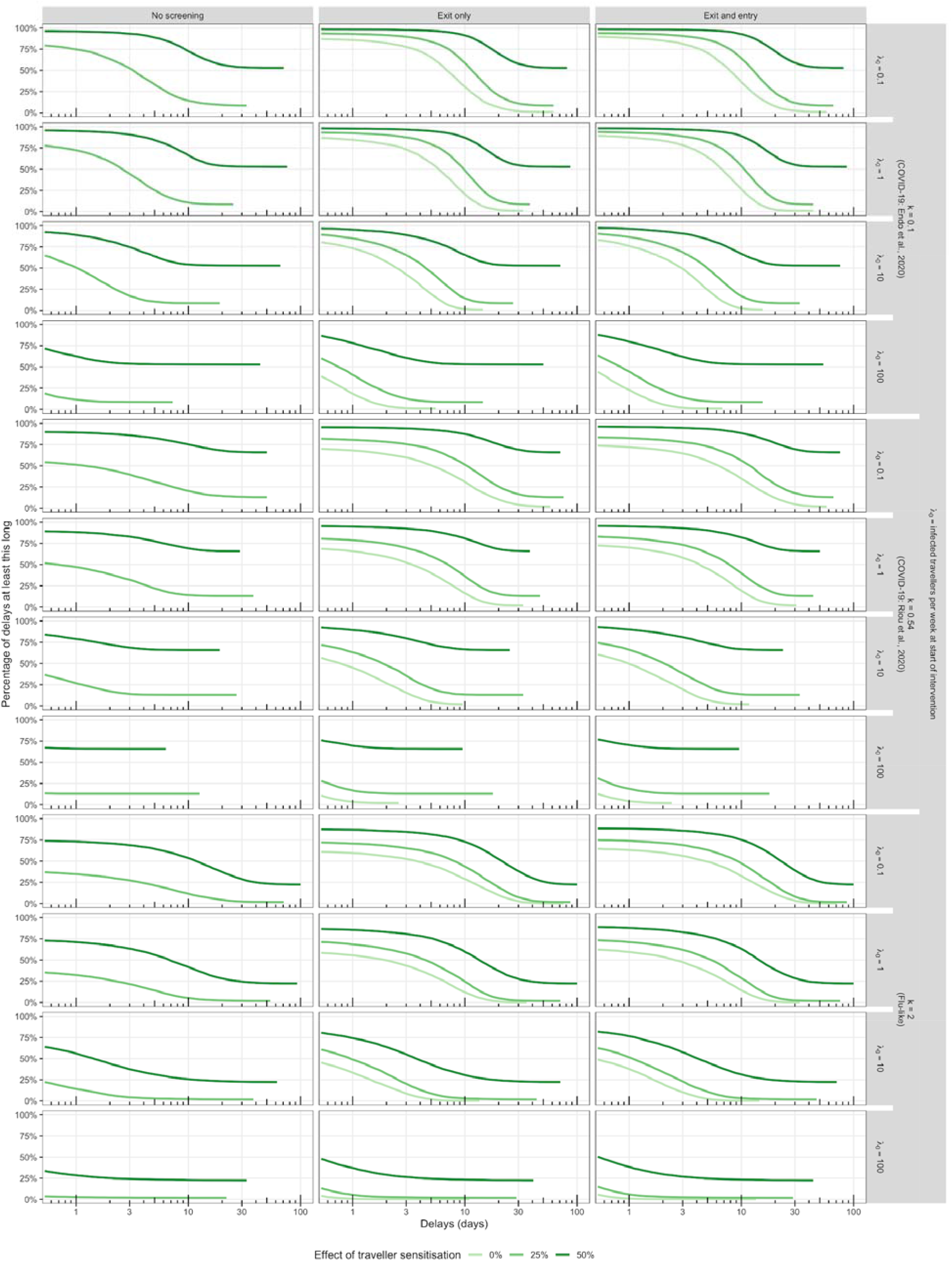
All scenarios for Figure 2 - complementary cumulative density functions (CCDF) of the estimated number of days an outbreak is delayed given an intervention consisting of a combination of traveller screening and sensitization and contact tracing. Within each panel, and for a given delay, the CCDF shows the percentage of simulations which result in a delay of at least that long for each level of traveller sensitisation (0%, 25%, 50%). Comparisons are made to no contact tracing and no screening.

**Figure S3:**
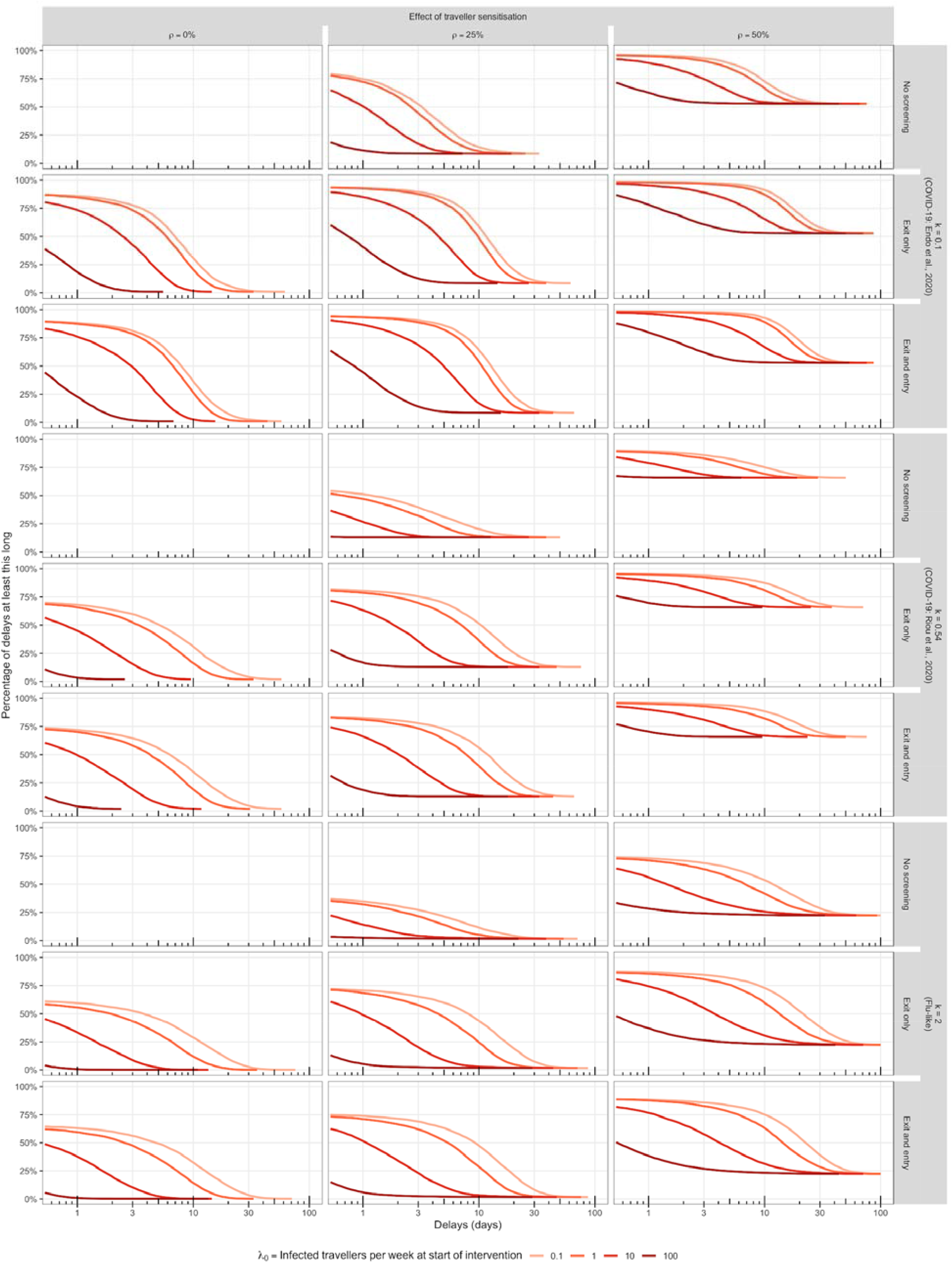
All scenarios for Figure 2 - complementary cumulative density functions (CCDF) of the estimated number of days an outbreak is delayed given an intervention consisting of a combination of traveller screening and sensitization and contact tracing. Within each panel, and for a given delay, the CCDF shows the percentage of simulations which result in a delay of at least that long for each rate of arrival of infected travellers at the time interventions are introduced (0.1, 1, 10 and 100 per week). Comparisons are made to no contact tracing and no screening.

**Figure S4:**
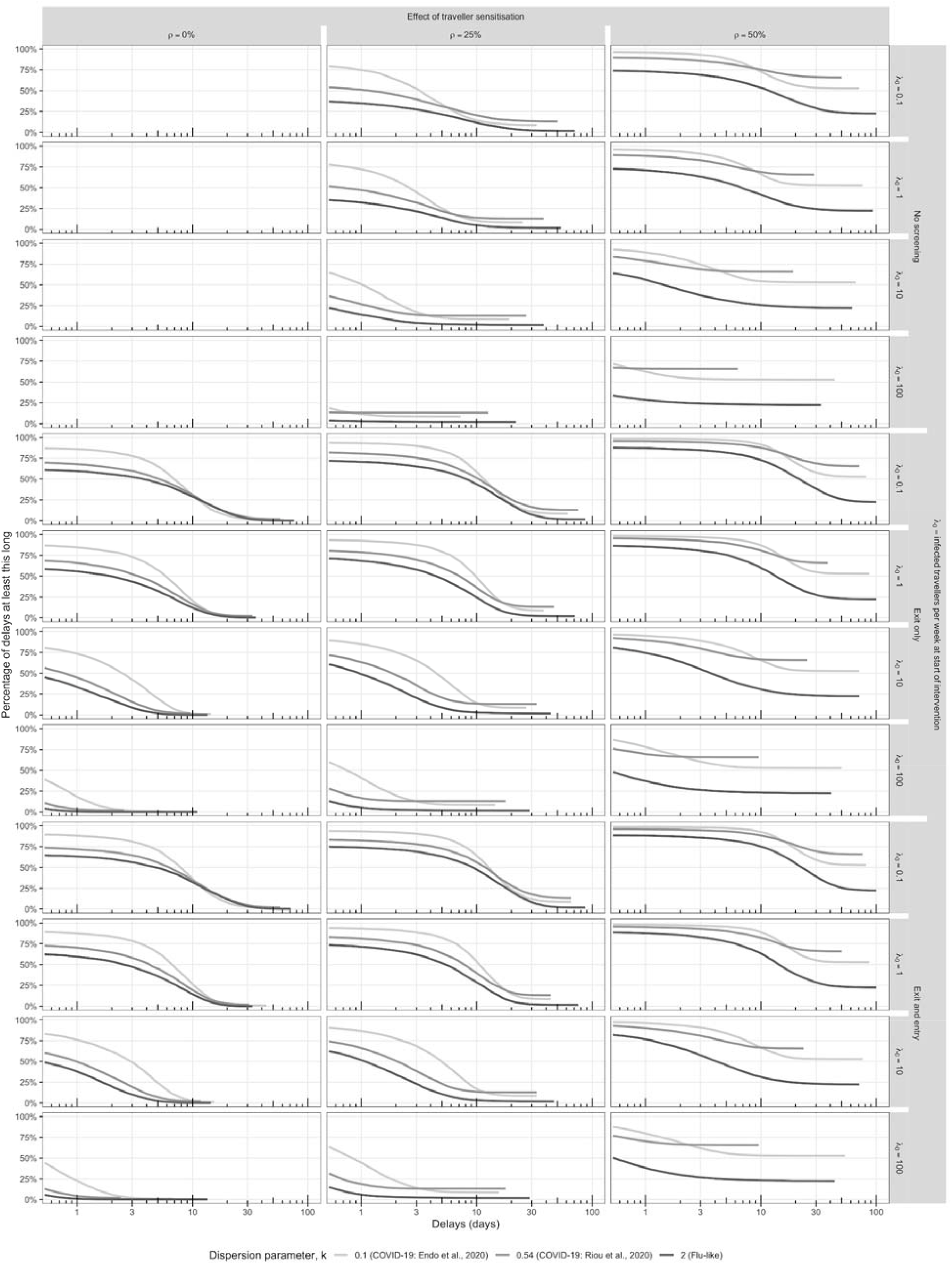
All scenarios for Figure 2 - complementary cumulative density functions (CCDF) of the estimated number of days an outbreak is delayed given an intervention consisting of a combination of traveller screening and sensitization and contact tracing. Within each panel, and for a given delay, the CCDF shows the percentage of simulations which result in a delay of at least that long for each dispersion parameter considered. Comparisons are made to no contact tracing and no screening.

Table S1 shows the percentage of the delays in Figures 1, S1-S3 that are infinitely long as a function of the traveller sensitisation. These values are independent of whether or not screening is performed, as well as *λ*_0_ and *k*.

**Table S1:**
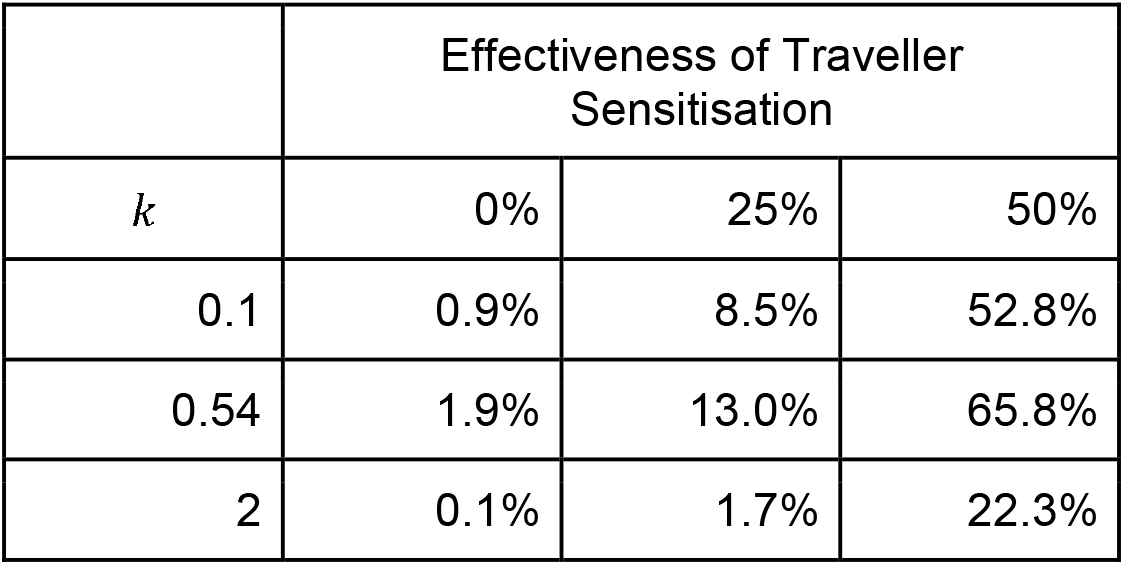
Percentage of delays which are infinitely long under varying levels of traveller sensitisation. Results from 5 x 10^3^ simulations.

|  | Effectiveness of Traveller Sensitisation |  |  |
| --- | --- | --- | --- |
| $k$ | 0% | 25% | 50% |
| 0.1 | 0.9% | 8.5% | 52.8% |
| 0.54 | 1.9% | 13.0% | 65.8% |
| 2 | 0.1% | 1.7% | 22.3% |

In Table S2 we present the full summary statistics of the sensitivity analysis.

**Table S2.**
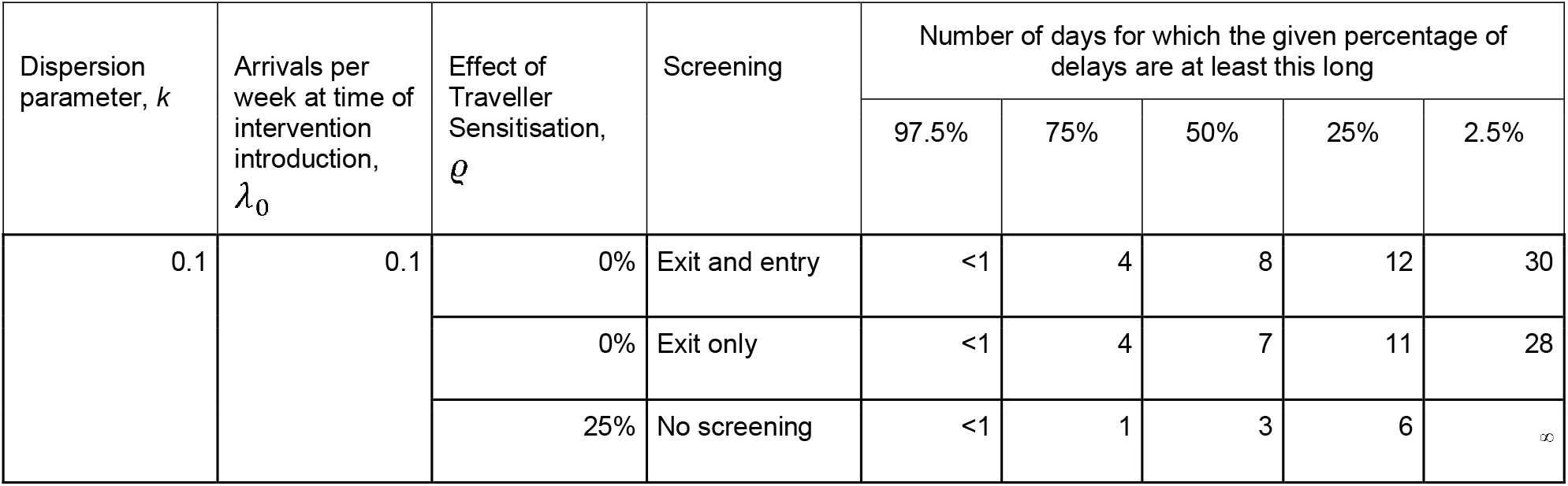
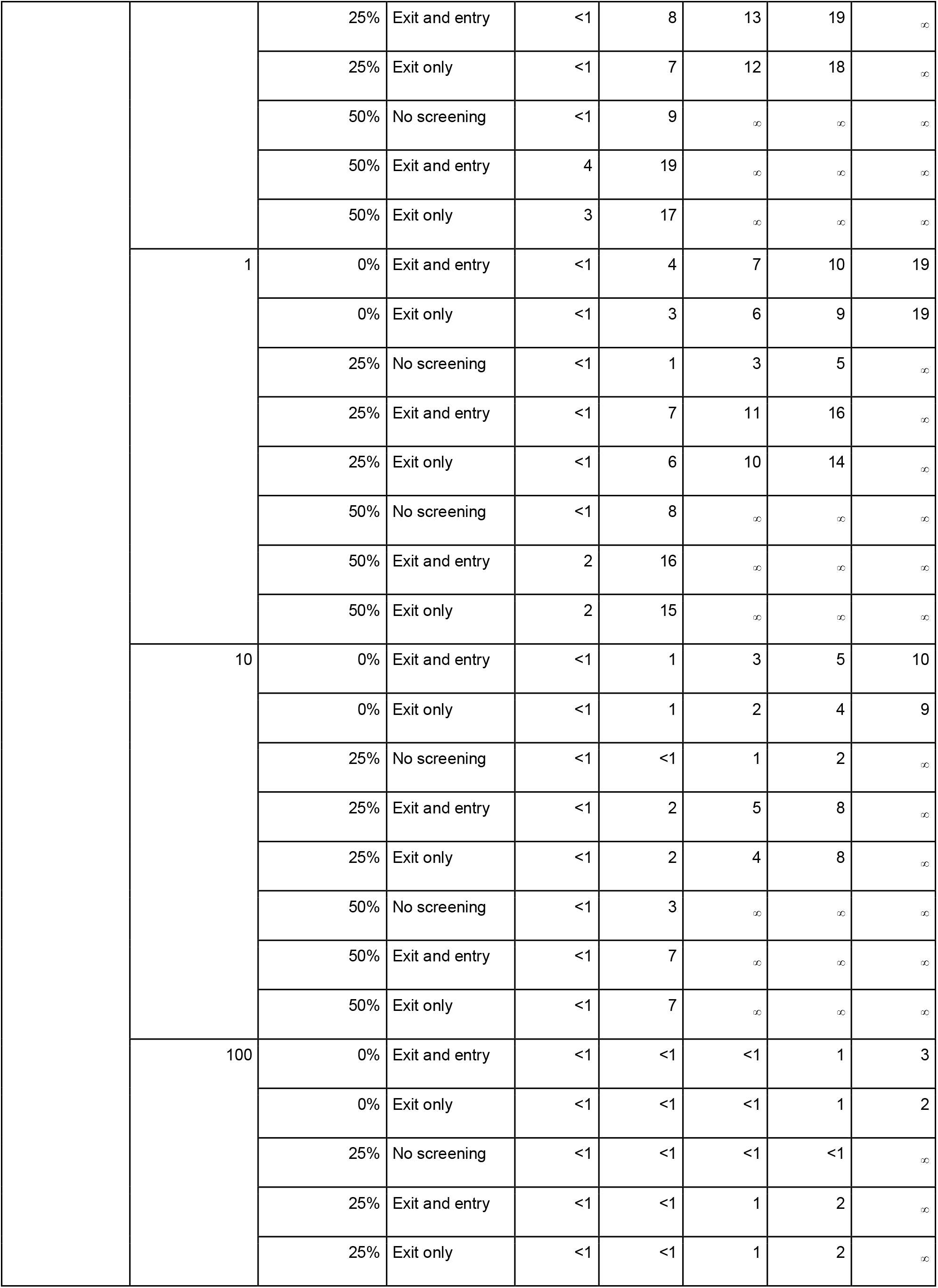
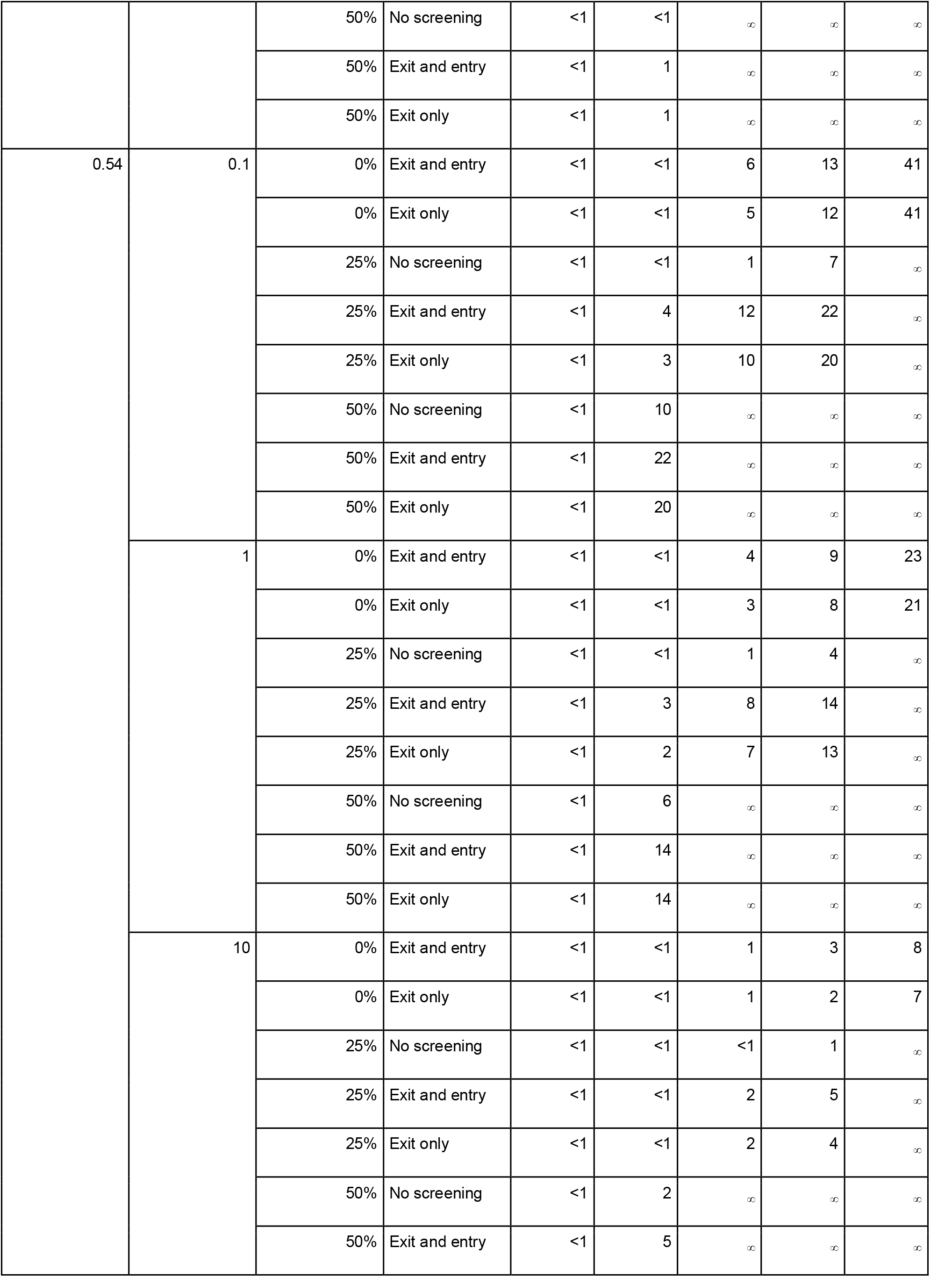
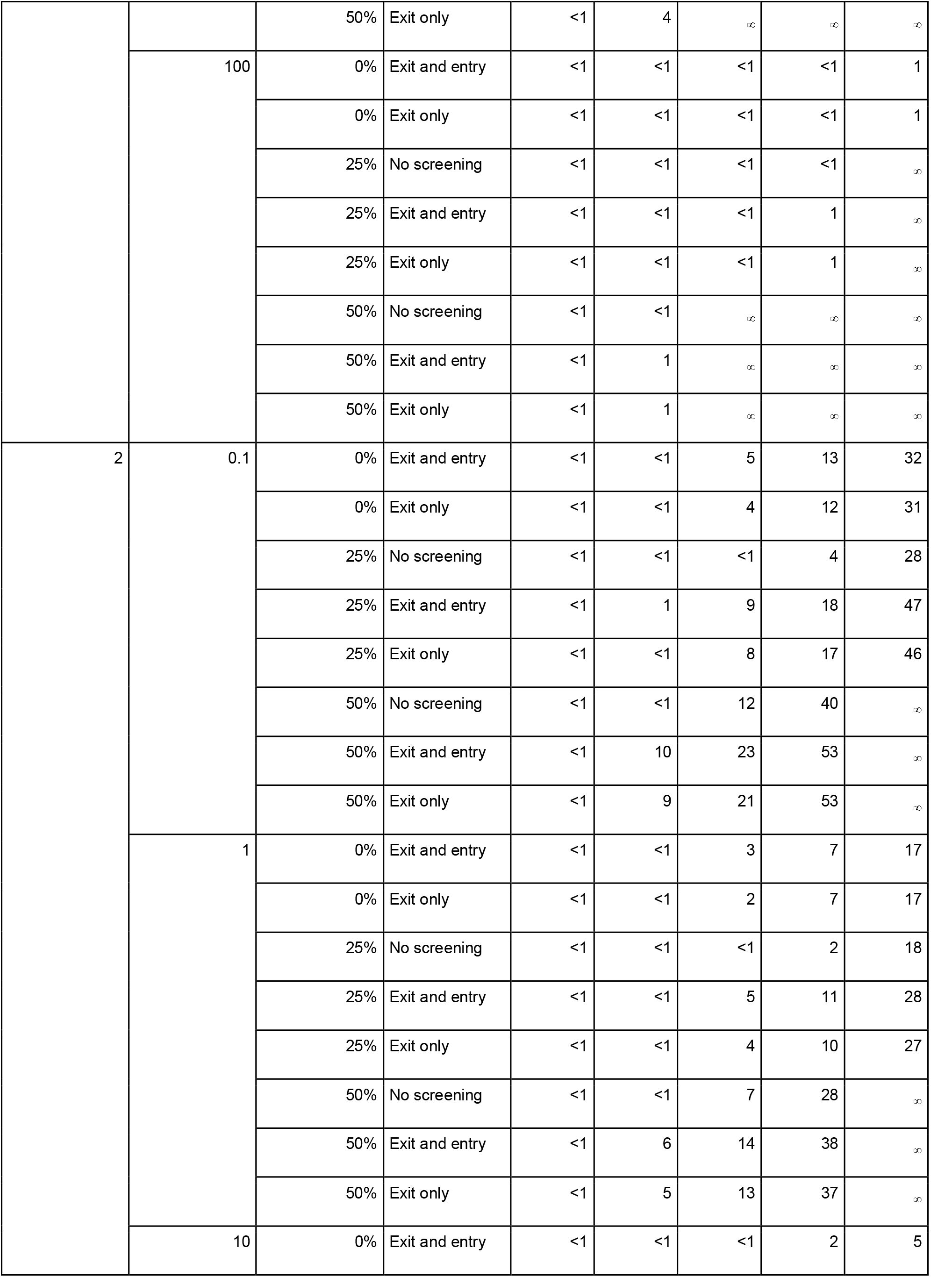
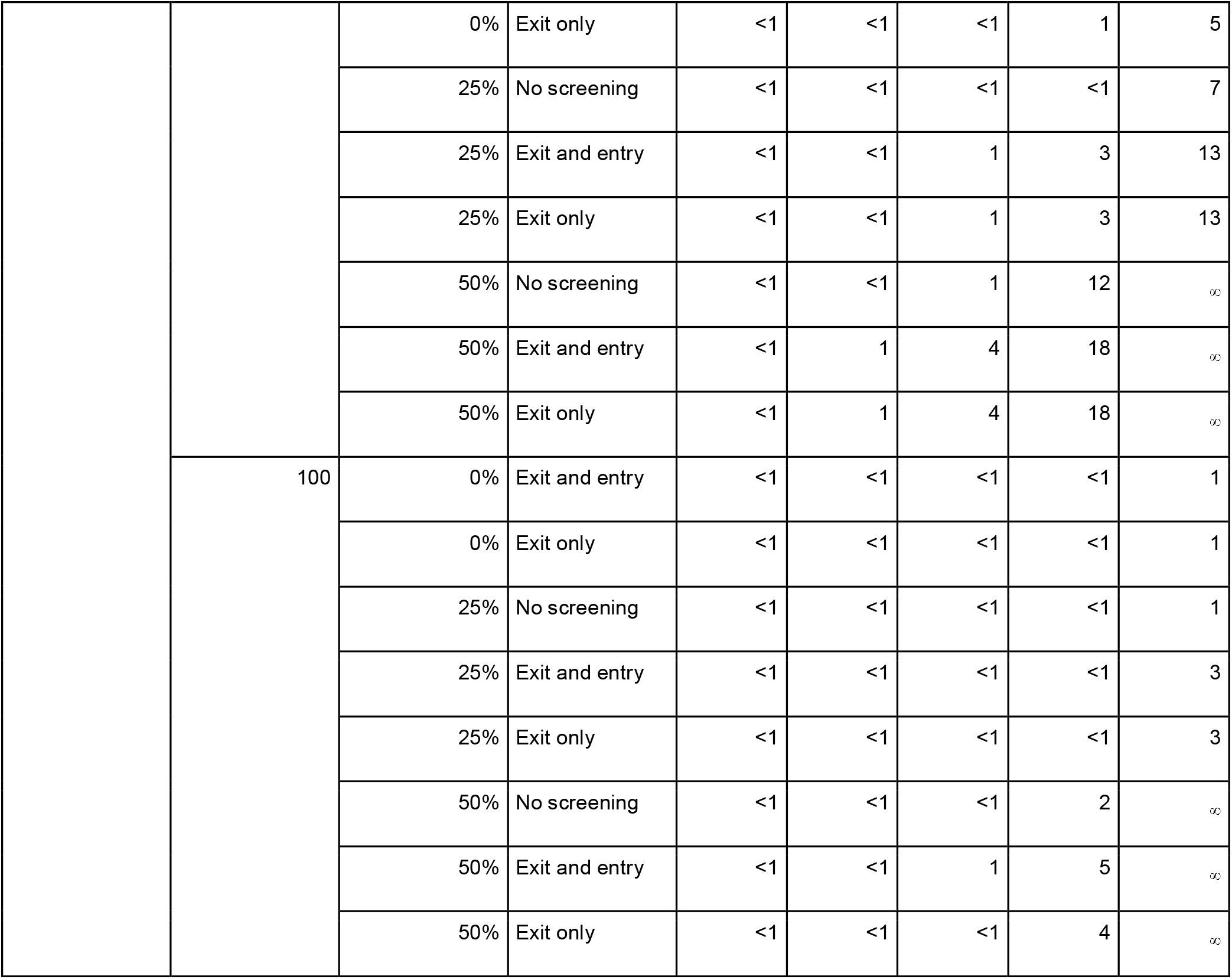
Sensitivity analysis summary statistics providing the inner 50% and 95% confidence intervals and medians (all rounded to the nearest day) for the estimated number of days an outbreak is delayed given an intervention consisting of a combination of traveller screening and sensitisation and contact tracing. Comparisons are made to no contact tracing and no screening (there are no “No screening” results at 0% sensitisation as this is the baseline against which comparisons are to be made).

